# COVID-19 serological survey using micro blood sampling

**DOI:** 10.1101/2020.10.09.20209858

**Authors:** Melissa M. Matthews, Tae Gyun Kim, Satoshi Shibata, Noriko Shibata, Christian Butcher, Jaekyung Hyun, Keon Young Kim, Theodore Robb, Siang Sheng Jheng, Masashi Narita, Tomoari Mori, Mary Collins, Matthias Wolf

## Abstract

During August 2020, we carried out a serological survey among students and employees at the Okinawa Institute of Science and Technology Graduate University (OIST), Japan, testing for the presence of antibodies against SARS-CoV-2, the causative agent of COVID-19. We used a FDA-authorized 2-step ELISA protocol (*1, 2*) in combination with at-home self-collection of blood samples using a custom low-cost finger prick-based capillary blood collection kit. Although our survey did not find any COVID-19 seropositive individuals among the OIST cohort, it reliably detected all positive control samples obtained from a local hospital and excluded all negatives controls. We found that high serum antibody titers can persist for at least up to 6.5 months post infection. Among our controls, we found strong cross-reactivity of antibodies in samples from a serum pool from two MERS patients in the anti-SARS-CoV-2-S ELISA. Here we show that a centralized ELISA in combination with patient-based capillary blood collection using as little as one drop of blood can reliably assess the seroprevalence among communities. Anonymous sample tracking and an integrated website created a stream-lined procedure. Major parts of the workflow were automated on a liquid handler, demonstrating scalability. We anticipate this concept to serve as a prototype for reliable serological testing among larger populations.

## Background and Introduction

At the beginning of 2020, COVID-19 emerged from Hubei province in southern China and quickly spread across the globe. Due to its proximity and direct flight connections between the epicenter in Wuhan and its capital Tokyo, the Japanese Nation was among the first to experience cases of the disease outside China, documenting its first case on January 15, 2020. In early April, as the tourism season in Okinawa began to ramp up, so did the new cases of COVID-19 in Okinawa. A first wave of COVID-19 cases peaked at the moderate rate of 10 cases/day in mid-April (total number 132 cases), which was effectively reduced by 5 weeks of lock-down measures to no additional cases within the next two months. From early August on, however, a second, much stronger wave of infections has, at the time of this report, culminated in more than 2,682 accumulated cases on the island (October 8, 2020). The present study was conducted at the beginning of this second wave.

Serological surveys which detect the presence of antibodies against SARS-CoV-2 antigens can provide important information to the government for issuing health care guidelines. At the beginning of April 2020, we obtained plasmids for coronavirus surface antigens from the Krammer Lab at the Icahn School of Medicine (NY, NY, USA). We established protein expression and purification in a mammalian cell line and set up an ELISA following the 2-step assay developed by the same group (*1*). Their assay has received emergency use authorization by the U.S. Food & Drug Administration (FDA) (*2*). We secured PCR-confirmed human sera from COVID-19 positive patients at the local hospital as well as negative controls from serum collected before December 2019. Once the assay itself was validated, we set up partially automated sample handling on a robotic liquid handler, established a website with a barcoding system for anonymous sample tracking, and conducted a serological survey of staff and students at our institution.

In emergency situations such as during COVID-19, the health care system is under stress; it cannot be expected that trained clinical personnel are available to draw patient blood by venous puncture. Furthermore, non-essential human traffic in hospitals and other health care institutions should ideally be limited to protect vital health care workers from risk of exposure to potential carriers of the virus. To overcome this limitation, we distributed easy-to-use, self-administered micro blood sampling kits to participants. The kit uses a single-use safety lancet to collect a few drops of capillary blood from the participant’s finger. We show that antibody titers obtained by micro blood sampling are equivalent to serum antibody titers from blood drawn by conventional venous puncture. This low-cost, easily deployable self-sampling method in combination with a highly sensitive and specific ELISA in a centralized testing lab provides a scalable solution that can enable serological surveys of larger populations.

## Methods

### Protein Expression and Purification

Plasmids for mammalian expression of the SARS-CoV-2 S (spike) protein receptor-binding domain (RBD, residues 319-541) and stabilized His-tagged SARS-CoV-2 S including a T4 foldon trimerization tag were generously provided by Florian Krammer and colleagues at the Icahn School of Medicine (NY, NY, USA) (*3*). Proteins were expressed in Expi293F cells and purified by Ni-NTA affinity and size-exclusion chromatography as described previously (*1*). The yield was 15 mg/L and 4 mg/L for RBD and the trimeric SARS-CoV-2 S, respectively. 500 µL aliquots of purified protein at a concentration of 10 µg/mL were frozen in liquid nitrogen and stored at −80 °C until use. We expressed and purified both the SΔcs (furin cleavage site deletion RRAR to A) and SΔcspp construct (cleavage site deletion and stabilizing mutations K986P and V987P) (*3*), but only the SΔcs protein was used in this study. Protein purity was verified by SDS-PAGE and confirmed by Western blot with MonoRab™ Anti-His Tag (C-term) Antibody (Nr. 25B6E11, GenScript, USA).

### Electron microscopy

The quality and folding of two proteins, the RBD and SARS-CoV-2 S, were verified by biochemical methods and correct folding and assembly of the trimeric spike protein was confirmed using cryo-electron microscopy (Figure 1). Sample solution was applied to a carbon coated copper grid and stained with 1 % uranyl acetate. The protein particles were visualized with a Talos L120 transmission electron microscope (Thermo Fisher Scientific, USA) operating at 120 kV acceleration voltage.

**Figure 1.**
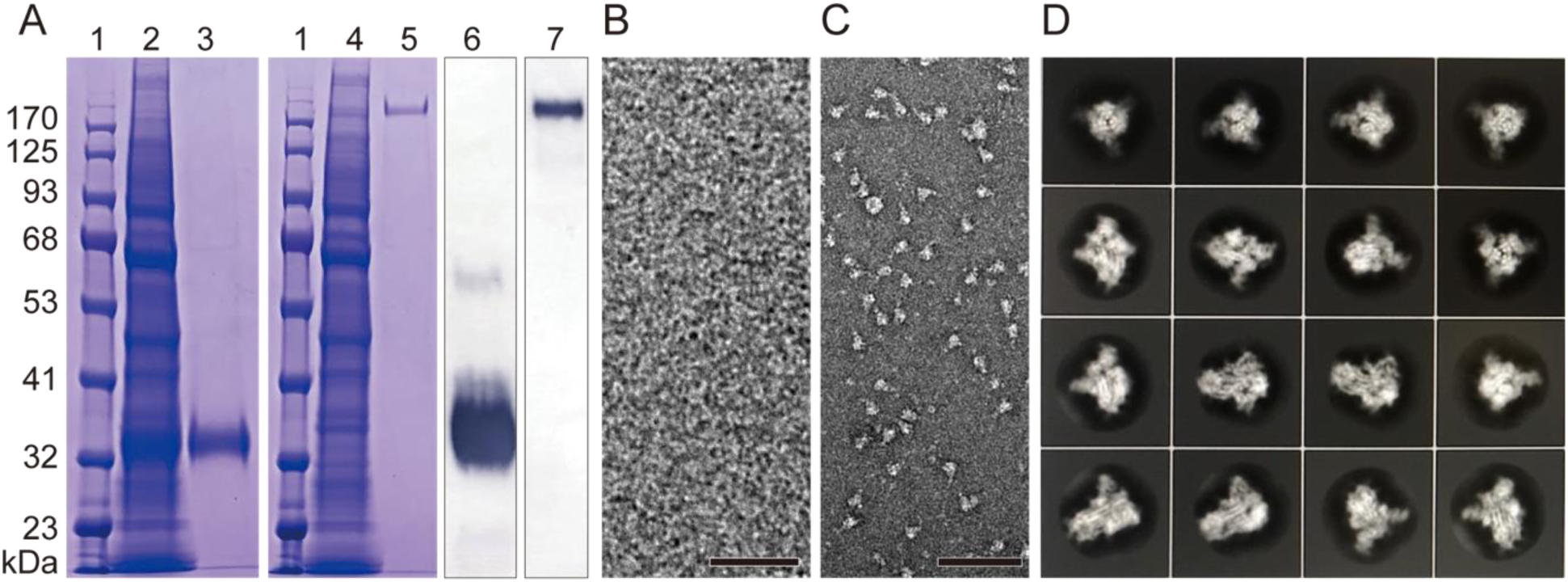
**A**. SDS-PAGE of expressed and purified RBD and spike proteins. Lane: 1, molecular weight marker; 2, cell lysate of expressed RBD; 3, purified RBD; 4, cell lysate of expressed SARS-CoV-2 S; 5, purified spike; 6 and 7, Western blot of purified RBD and spike, respectively. **B** and **C**. Electron micrographs of negatively stained purified RBD and spike protein, respectively. **D**. 2D class averages from boxed aligned single-particle cryo-EM images of the trimeric spike protein, showing secondary structure elements indicative of proper folding. Box size 28 nm. Scale bars 20 nm 100 nm in B and C, respectively. Original images are available as supplementary data files.

For cryo-EM, 3 µL of purified trimeric spike sample solution was applied to UltrAuFoil R1.2/1.3 grids pre-treated with a Solarus advanced plasma system (Model 950, Gatan, USA) for 60 seconds at 25 °C in a 23 % H_2_, 77 % O_2_ gas mix. The grids were blotted and vitrified in a Vitrobot Mark IV (Thermo Fisher Scientific, USA) using a liquid ethane-propane mixture. Cryo-EM grids and particle density were optimized by screening based on protein concentration in the range of 2-3 mg/mL. Particles Images of trimeric spike protein in amorphous ice were collected with a Titan Krios transmission electron microscope (Thermo Fisher Scientific, US) operating at 300 kV on a Falcon-3EC camera (Thermo Fisher Scientific, US) in counting mode, at a pixel size of 1.08 Å (at the specimen level). Frames from 1,479 movies were aligned, dose-weighted and summed using motioncor2 (*4*). Contrast transfer function (CTF) estimation, particle picking and 2D classification were performed with RELION 3.1 (*5*). A total of 1,119,504 particles were picked semi-automatically, of which 297,144 particles were retained after 2D classification.

### Sample collection and heat inactivation

The micro blood sampling kits were assembled in 8×12 cm sealable plastic bags, including BD Microtainer^®^ contact-activated lancet (Becton Dickison, NJ, USA), blood collection tube containing a coagulant and a separator gel (Greiner Bio-one MiniCollect^®^ TUBE 0.5/0.8 CAT Serum Separation Clot Activator gold cap), packaged alcohol wipes, adhesive bandage, and peelable sticker with a linear tube barcode (Code 128) and a matching QR code encoding a link to our website. After finger prick with the safety lancet, users collected 1-5 drops (approx. 30-150 µL) of capillary blood into the serum separation tube and attached the linear barcode. Serum separation tubes were centrifuged at 5,000×g for 5 min at 4 °C under biosafety level 2 (BSL2) conditions, then inactivated in a water bath at 56 °C for 1 hr. Heat-inactivated samples were stored at 4 °C until use for ELISA. After scanning tube barcodes for each plate into a CSV file with a hand-held scanner, separated serum was transferred into barcoded 96-well plates using manual pipetting.

### ELISA and automation

We followed the 2-step ELISA protocol developed by the Krammer lab (*1*). ELISA plate formats were modified as depicted in Supplementary Figure 1C. Modified plate designs included two dilution series of positive control, which provided an internal standard. Additional negative controls were included to provide a cutoff that was consistently within the suggested range of 0.15-0.2 AU (Absorbance Units) at 492 nm.

We implemented a partially automated workflow on a Beckman Biomek FX^P^ (Beckman Coulter, Indianapolis, IN, USA) liquid handling robot for two steps of the ELISA protocol, namely, for (i) dilution and transfer of serum samples for RBD screening plates and for (ii) plate developing and reading of all plates. Plates were stacked to maximize throughput and usage of available deck space. Automated dilution and transfer of serum samples utilized a 96-well plate containing 5X diluted serum in PBS, termed the D1 (dilution 1) plate as a source plate. Sample from the D1 plate was diluted with a PBS-T milk solution in a second 96-well plate, termed the D2 (dilution 2) plate, and finally transferred to the destination ELISA plate according to the published protocol (*1*). After dilution, D1 plates were immediately wrapped in parafilm and stored at 4 °C. If a given sample tested positive against RBD antigen in step 1, a fresh 5x diluted sample was prepared using solution remaining in the serum separation tubes. If the volume of the remaining serum was insufficient, the 5x diluted sample from the D1 plate was used for the confirmatory step 2.

For automated plate development and reading, 100 µL of Sigma*Fast*™ OPD development solution (Sigma Aldrich) was prepared according to the described protocol and added to all wells. A wait time was programmed in the liquid handler workflow so that exactly 10 minutes after addition of OPD, 50 µL of 3 M HCl were added to quench the reaction. Immediately after addition of HCl to a given plate, the plate was automatically loaded into and read on a DTX800 multimode plate reader (Beckman Coulter, Indianapolis, IN, USA) at 492 nm wavelength. Functions were added within the automation code to automatically calculate threshold values for each plate and output a “positive” or “negative” result for each sample. All other steps, including washing on an AquaMax 4000 plate washer (Molecular Devices LLC, San Jose, CA, USA), were carried out manually.

### Controls and Standards

Three positive controls in the form of intravenous blood sera from confirmed SARS-CoV-2 PCR-positive patients (collected 10-30 days post onset of symptoms) were obtained from Naha Municipal Hospital, Naha, Japan. After titer analysis of all three samples (data not shown), the two samples with the strongest titers were pooled and used as the positive control for all assays. Positive controls (collected at least 90 days after onset of symptoms) for validation of the capillary blood collection method were obtained from Okinawa Chubu Hospital, Uruma City, Japan. Negative controls taken from patients prior to November 2019 were obtained from Naha Municipal Hospital from intravenous blood, and from a commercial serum pool (Human Serum from human male AB plasma, Sigma Aldrich H4522-100ML, Batch #SLCD1948, serum was pooled prior to August 2019). Human MERS-convalescent serum and SARS-CoV-2 convalescent plasma (NIBSC code 20/130) were obtained from the National Institute of Biological Standards and Control, UK.

### Calculation of thresholds

The threshold for each step 1 plate was defined as the average of the negative controls plus 3 standards deviations of the negative controls as described previously (*1*). The average and median threshold for all step 1 plates was 0.161 AU and 0.166 AU, respectively. Initially, the threshold for the step 2 plate was calculated in the same manner as step 1. However, the threshold calculated with only three negative controls was below the recommended range (0.15-0.2 AU at 492 nm). Therefore, another threshold, calculated as 4-times the average blank, which has been demonstrated to be valid for identifying anti-SARS-CoV-2 antibody-positive samples was used instead (*6*). Average and median threshold for all step 2 plates were 0.196 AU and 0.1868 AU, respectively.

### Calculation of predictivity values

The positive predictivity value (PPV) describes the likelihood that a sample which tests positive is a true positive. The negative predictivity value (NPV) describes the likelihood that a sample which tests negative is a true negative (*7*). Estimated predictivity values were calculated using the following equations:

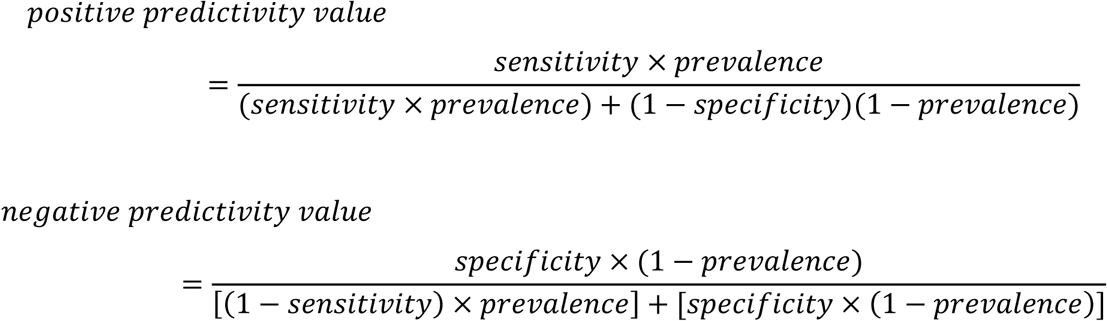

### Website-based platform for test application and anonymous results reporting

Test results were disseminated using a custom web application developed by the OIST IT section and deployed through Microsoft Azure cloud. The front page (Supplementary Figure 1B) contained a disclaimer, an instructional video (Supplementary Information), a printable instruction sheet, and other basic information for participants.

The QR code printed and included in each blood sample collection kit was encoded with a unique web URL corresponding to the sample ID of the sticker placed on the sample tube (Supplementary Figure 1A). The stickers and QR codes were created with the mailing feature in Microsoft Office 365 Excel. Following a simple user flow (Supplementary Figure 2), participants could check their test results anonymously either by scanning the QR code with a smartphone or by entering the 6-digit sample ID matching the linear tube barcode.

**Figure 2:**
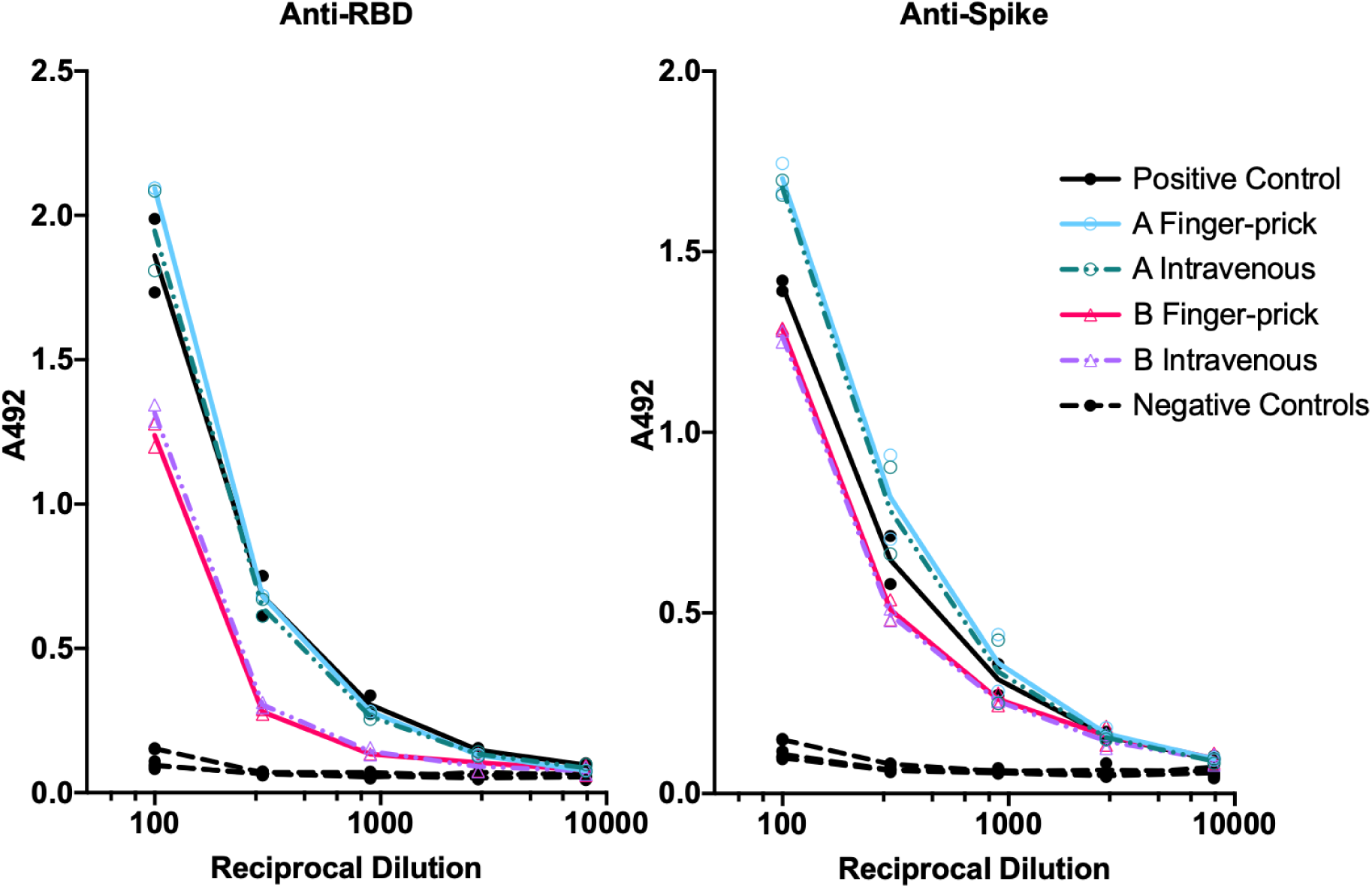
Serum antibody titers from intravenously collected blood and capillary blood obtained by finger prick are equivalent. Individuals A and B provided blood samples both intravenously and also by micro-blood collection at least 90 days post exposure. Their endpoint titers were comparable to the positive controls. Each sample was tested in duplicate on the same ELISA plate, either coated with SARS-CoV-2 spike RBD (left) or with SARS-CoV-2 trimeric spike protein (right). Each replicate is plotted. Line connects average values. Positive control was pooled serum collected from two SARS-CoV-2 PCR-positive individuals 10-30 days after onset of symptoms. Negative controls were collected from individuals prior to November 2019.

Upon inputting a valid sample ID, participants were prompted to accept a disclaimer before viewing results. After accepting the disclaimer, participants were able to retrieve the status of their sample, including guidance on interpreting test results. Optional information about gender, prior symptoms, age, travel history could be entered voluntarily. This metadata was collected anonymously through a survey on the SurveyMonkey (https://www.surveymonkey.com) platform and linked to the sample ID which was passed as a hidden field via the participant’s web browser. Each sample ID was stored in a database table along with values representing test results and disclaimer acceptance. The web application queried this database to retrieve the status of the requested sample ID. The sample IDs were composed of six characters generated randomly from the set of alphanumeric characters (a-zA-Z0-9) (random string generator https://www.random.org/strings/), excluding letters O, I, l, Z, Q and numbers 0, 1, 2, 9 to minimize human read errors. Out of approximately 10^10^ combinations, 1000 unique IDs were randomly selected. Some additional security features, such as request rate limiting, were implemented to restrict the use of automated tools to retrieve results for all possible sample IDs. Basic functions were developed to allow batch import and export of test results in a delimited text format, allowing easy transfer of data between the testing system and the web application database.

### Sequence alignments

The full-length sequences of SARS-CoV, MERS-CoV S protein (UniProt ID: P59594 and K9N5Q8, respectively) were aligned pairwise using ClustalW2 (*8*), against the SΔcs sequence (*3*) (Supplementary Figure 3).

**Figure 3:**
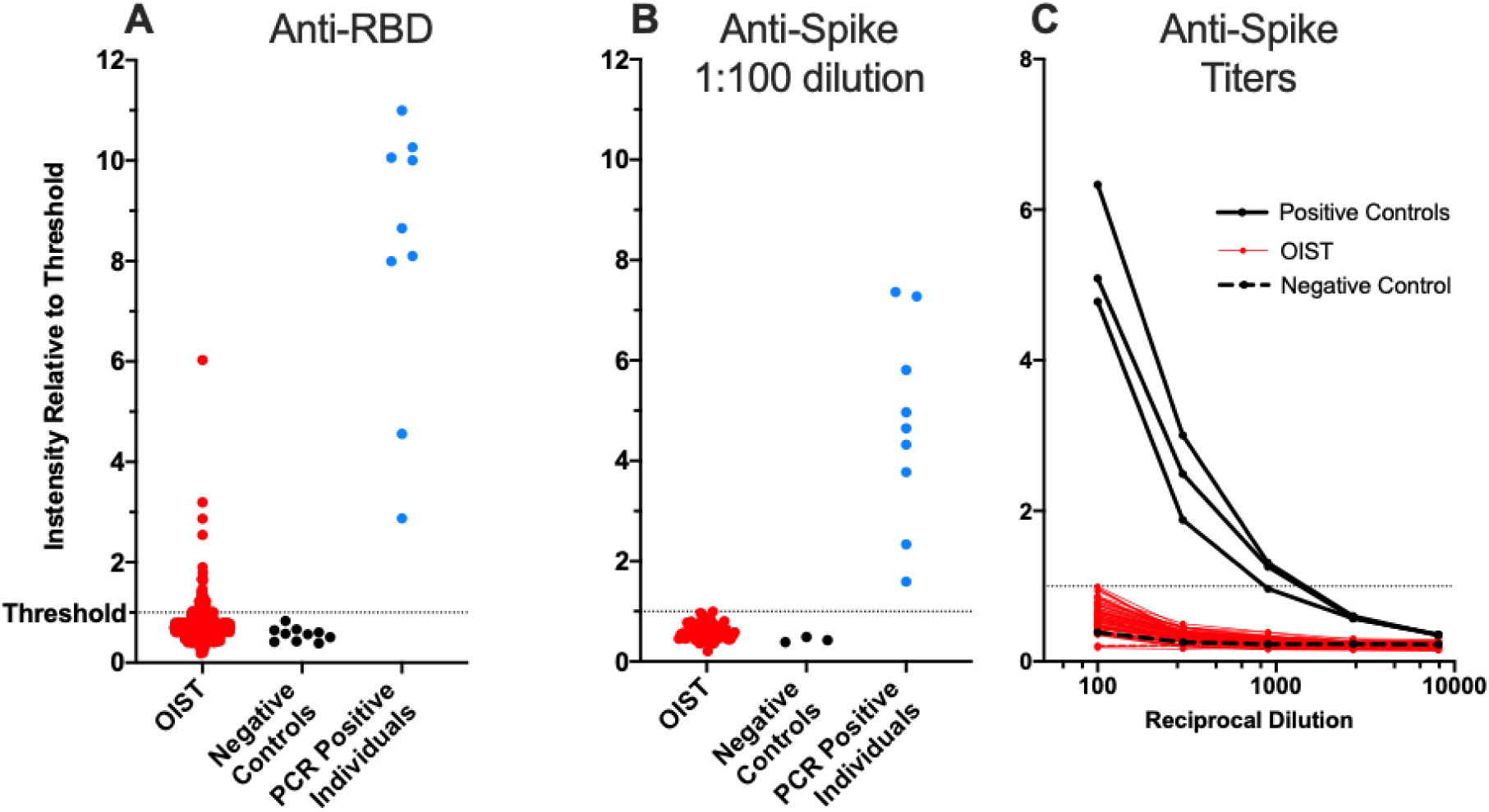
Summary of OIST Serological Survey. **A.** In assay step 1 (anti-RBD ELISA), 63 OIST samples were above the threshold (3 standard deviations above the average negative control). **B**. All positives from step 1 were tested in step 2 (anti-spike ELISA). No OIST samples were above the threshold (4 times the average background). **C**. Step-2 titers were measured at dilutions 1:100, 1:300, 1:900, 1:2700, and 1:8100. Positive controls showed much stronger signal than the OIST samples even at high dilution. The dotted line indicates the respective threshold level. All negative controls are samples taken from patients prior to November 2019. Sera from PCR positive individuals were collected at least 10-30 days post onset of symptoms.

### Figure preparation and digital processing

Images of protein gel and Western blot in Figure 1A were acquired by smartphone camera, cropped and adjusted for intensity level in Adobe Photoshop 2020 (Adobe Inc, USA). Original unaltered gel images are available as supplementary files. Original uncropped digital electron micrographs used for Figure 1B and 1C are available as supplementary files. Graphs in Figures 2-5 were plotted with GraphPad Prism v8.4.3 (GraphPad Software, USA), using the scatter plot function (Figure 2).

## Results

### Protein preparation and validation

SARS-CoV-2 trimeric spike and its receptor-binding domain (RBD) were expressed in mammalian cells and purified. The quality was verified by Western blot and electron microscopy (Figure 1 A-C). We performed single particle cryo-EM and 2D-image classification. This confirmed that the trimeric spike protein was properly folded and assembled (Figure 1D).

### Validation of micro blood sampling method

Serum samples are typically separated from intravenously collected blood. To confirm that the finger-prick method neither causes unforeseen complications nor affects assay sensitivity, blood samples from two confirmed SARS-CoV-2 PCR-positive individuals were taken both intravenously and by finger-prick on the same day. Although these samples were collected at least 92 days post exposure, they retained a high antibody titer comparable to positive controls, which were collected 10-30 days post onset of symptoms.

The MiniCollect^®^ capillary blood collection tube contains a prefilled volume of a polymer gel with a lithium heparin coagulant, which induces blood clotting and can separate the blood clot from serum by centrifugation. Separated serum from four samples were tested in a dilution series in duplicate on both an RBD-coated ELISA plate and a trimeric spike-coated ELISA plate according to the “step 2” ELISA protocol (*1*) (see Methods and modified plate layout in Supplementary Figure 1C). For both individuals, titers of intravenous and capillary blood were similar on both the RBD-coated and the spike-coated plate (Figure 2).

### Serological Survey

Overall, 675 sample tubes were collected and processed. Among all the samples received, zero samples showed signal above threshold in both the RBD screening plate and the spike confirmatory plate.

Samples were typically collected, serum separated, and heat-inactivated at the end of each day. Nonetheless, most blood samples can be stored at room temperature in the serum separation tube for several days, in some cases with larger blood volume for up to 1 week. Longer than 1 week is not recommended because the blood begins to dry up. Antibody titers after serum separation have been reported as stable for up to 6 weeks when stored at 4 °C (*9*). Some participants had difficulty collecting their blood by themselves. In such cases, we encouraged participants to visit the nurses in our institute’s Health Center for assistance.

The results for RBD ELISA step 1 are summarized in Figure 3A. Intensities were scaled with respect to the threshold for each plate. Results were compared with multiple negative controls and a collection of serum samples from SARS-CoV-2 PCR positive individuals. A subset of 63 OIST samples had serum antibody reactivity above the established threshold, and only a few were at the same level as the PCR-positive individuals. Titers from all PCR-positive individuals were above the threshold. Samples from all PCR-positive individuals were taken by capillary blood collection at least one month post onset of symptoms.

Results for the final ELISA step 2 are shown in Figure 3B and 3C. Of the 63 OIST samples that were positive in RBD step 1, all had serum antibody reactivity below threshold for all 5 dilutions when tested against the SARS-CoV-2 trimeric spike protein in step 2, indicating a 9.9% false positive rate in the highly sensitive step 1. Antibody titers (Figure 3C) of serum samples from all PCR positive individuals were above the threshold for at least two dilutions. Results were classified as “positive” (above threshold), “negative” (below threshold), or “undetermined”. All RBD-positive OIST samples were below threshold in the anti-spike ELISA (Figure 3B and 3C). The most common cause of an undetermined result was the failure to provide sufficient serum. This was the case for 40 of the 675 samples that we received. If the result was undetermined, the participant was encouraged to pick up another kit and try again.

From the SurveyMonkey platform, 206 completed surveys were received, or 31 % of all samples collected. The collective results showed that 17 % of the survey takers believed that they had experienced some COVID-19 symptoms in the past 6 months and 31 % had travelled outside of Okinawa in the past 6 months. The age distribution was as follows: 20-40 years old, 60 %; 40-60 years, 36 %; greater than 60 years, 4 %. The gender distribution was roughly equal.

### Assay Specificity

The specificity of the Mount Sinai Hospital Clinical Laboratory COVID-19 ELISA Antibody Test has been reported as 100 % for 74 negative control samples (*10*) and the developers demonstrated no cross-reactivity against the common coronavirus strain NL63 (*3*). Of note, as part of the protocol establishing process, we used a SARS-CoV-2 convalescent plasma (NIBSC code 20/130) and human MERS-convalescent serum to confirm the specificity of the assay in both steps. As expected, SARS-CoV-2 convalescent plasma showed strong reactivity in both plates. MERS-convalescent serum, on the other hand, was negative in anti-RBD cross-reactivity, but positive in anti-spike cross-reactivity (Figure 4).

**Figure 4:**
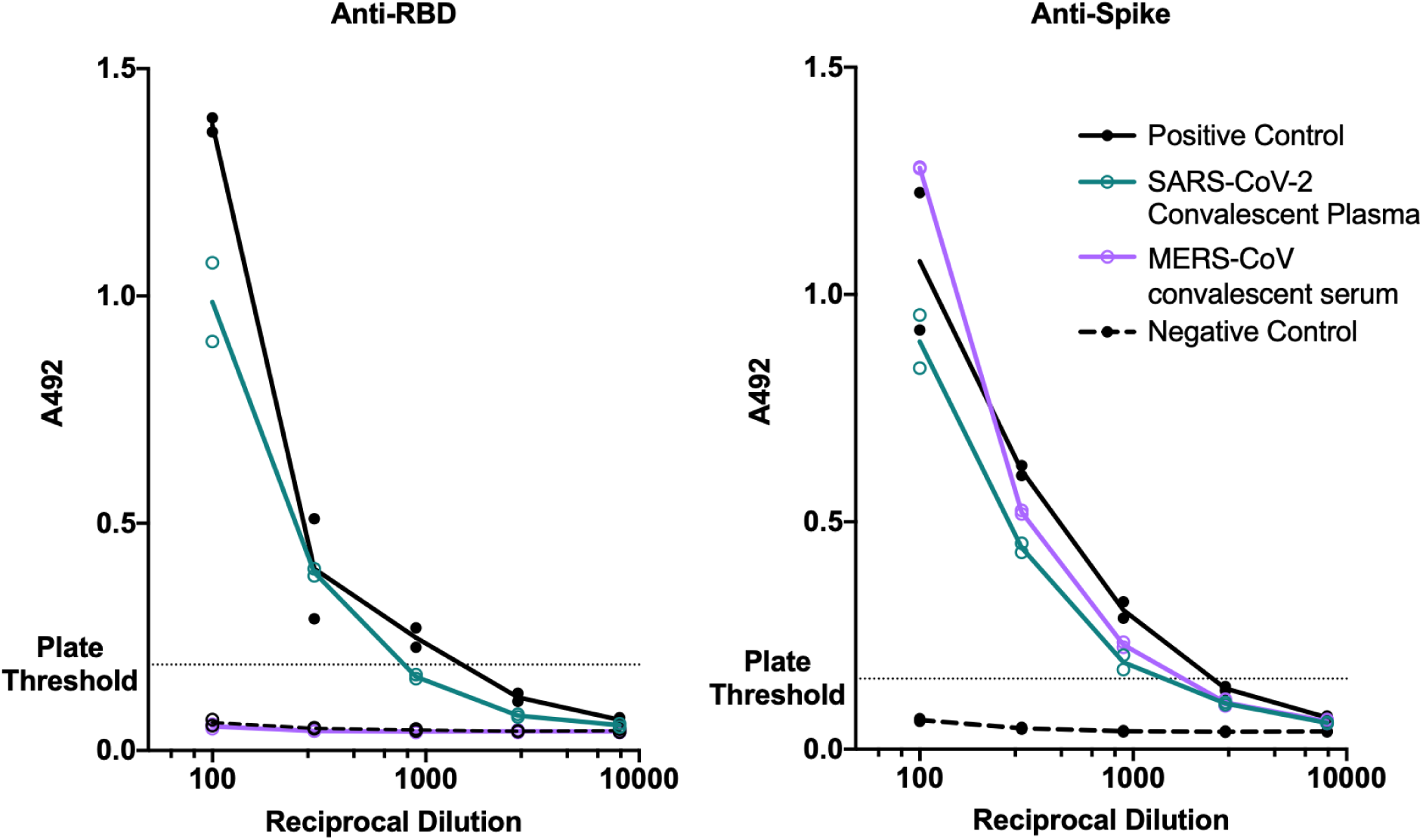
Titers of SARS-CoV-2 and MERS-CoV convalescent serum in RBD and spike ELISA. Each sample was tested in duplicate on the same plate. Line connects average values. Positive control is pooled serum from two CoV-2 PCR positive individuals 10-30 days after onset of symptoms. Negative control is from a commercial serum pool.

To understand the basis of the MERS-convalescent serum cross-reactivity, we performed an amino acid sequence alignment of SARS-CoV-2 spike and MERS-CoV spike protein. Sequence alignment demonstrated that the SARS-CoV-2 RBD shares 18.7 % identity with MERS-CoV RBD, but alignment of the full-length spike sequence demonstrated higher 32.3 % sequence identity between SARS-CoV-2, and MERS-CoV (Supplementary Figure 3).

### Longevity of Antibody Titers

We obtained multiple samples from two PCR-confirmed SARS-CoV2 positive individuals over an extended period of time and analyzed their serum antibody titers against the spike protein by ELISA. Individual 2 has a known exposure date, because the person was part of a documented cluster infection. Antibody titer was measured by anti-spike ELISA in a series of dilutions (Figure 5) in quadruplicate. Anti-spike antibodies titer remained above threshold 6.5 months post exposure.

**Figure 5:**
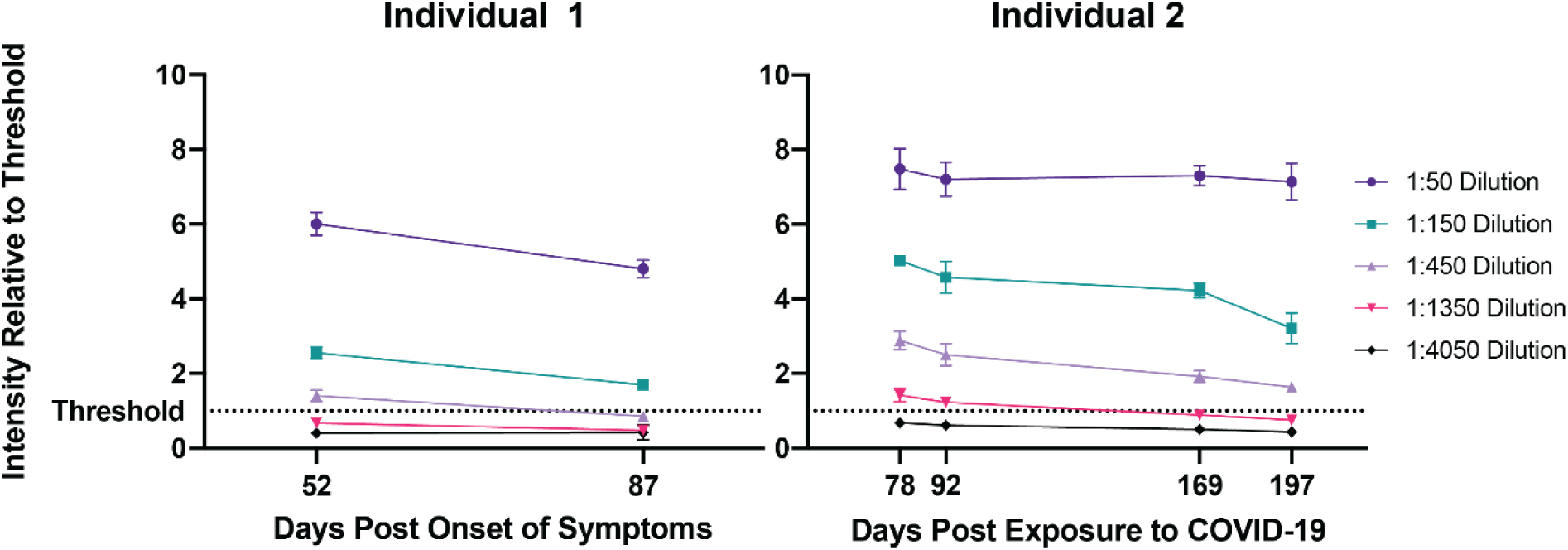
Resiliency of anti-spike antibody titer over time. Serum samples from two recovered COVID-19 patients were collected at 52 and 87 days post onset of symptoms (Individual 1, left) or at 78, 92, 169, and 197 days post exposure to the COVID-19 virus (Individual 2, right). Anti-spike antibody titers were measured by ELISA at 50, 150, 450, 1350, and 4050 times dilution in quadruplicate. The average and standard deviation for each dilution of each sample is plotted, revealing detectable antibody titers at 1:450 dilution for up to 6.5 months post exposure.

## Discussion

### Micro blood sampling is a good alternative to venous blood collection

Micro blood sampling by finger prick at home avoids unnecessary risk and effort by healthcare workers and allows parallel collection of large numbers of samples within a short period of time. The combination of patient-driven sample self-collection with a standardized highly specific and selective ELISA in a centralized lab improves the reliability of results while keeping costs low. Serum can be separated from whole blood up to 1 week after collection. Provided that temperatures are kept close to ambient temperature, this would allow for shipping of samples from collection site to the testing site by mail. The main bottleneck is serum transfer from collection tube to ELISA plate. This step may be accelerated with a sophisticated integrated liquid handling system. We showed that antibody titers from capillary blood serum are equivalent to titers measured from venous blood (Figure 2). The Infectious Diseases Society of America Guidelines on the Diagnosis of COVID-19 currently make no recommendations for or against the use of capillary blood in serological assays due to a knowledge gap on the subject (*11*). We hope that the success of our study begins to fill this knowledge gap, paving the way for further proof-of-concept studies using capillary blood. Our survey revealed that micro blood sampling of capillary blood is a practical, cost-efficient method using as little as one drop of blood (approximately 30 µL) per sample. Such a small sample volume would even allow testing of newborns using neonatal heel prick.

### Anonymization and web-based reporting

The web-based platform served as an efficient means to conduct this study anonymously while providing instructions and communicating results to participants. Anonymization was achieved by randomly barcoded samples without the need to enter personal data. From the 1,154 staff and students at OIST (as of May 2020), 675 samples were received, of which only 41 samples (6.1%) could not be tested due to lack of serum or improper sample collection. The success of our practical web-based method indicates that this method may be used for larger epidemiological studies.

### Significance of OIST results

The serological survey of students and employees carried out at OIST on Okinawa, Japan, in August 2020 revealed no seroconversion among this small population. The test has reliably identified all previously confirmed PCR-positive individuals and controls, with reactivities as high as 7 times threshold. Our results assert strong confidence in this 2-step assay, which has received FDA-emergency approval (*2*).

Okinawa has, thus far, enjoyed moderately low numbers of COVID-19 cases among its population. Other recent serological surveys in Japan, have found seroprevalence as low as 0.43 % (among 44,066 employees and business partners of the company Softbank and healthcare workers across Japan) (*12*), 0.1 % (1,971 citizens of Tokyo), 0.03 % (3,009 citizens in Miyagi Prefecture) and 0.17 % (2,970 citizens of Osaka prefecture) (13). At such low seroprevalence, high assay specificity is critical to achieving a high predictive value (*14, 15*). Although accurate estimation of the actual seroprevalence among OIST staff and students (including the 45 % of employees that did not participate in the survey) is not possible, assuming a seroprevalence between 0.03 % (Tokyo) and 0.43 % (Softbank), with a known assay specificity of close to 100 % and a sensitivity of 92.5 % for the Mt. Sinai Antibody Test used here (*10*), we estimate the negative predictivity value of our assay to be greater than 99.9%.

### Longevity of antibody titers

Analysis of serum antibody titers against SARS-CoV2 spike protein taken at multiple time points from two PCR-positive individuals post onset of symptoms (or post verified exposure) revealed only moderate decrease over time, with robust antibody titers above threshold persisting for up to 6.5 months (Figure 5). In combination with recent reports of high efficacy of vaccines currently in phase-3 trials, our results add to the growing amount of evidence that spike-based vaccines may be able to elicit longer-lasting immunity.

### Cross-reactivity of MERS convalescent serum with SARS-CoV-2 spike protein

SARS-CoV, SARS-CoV-2 and other human coronaviruses such as NL63 use aceE2 as receptor (*16, 17*) mediated by the class-1 fusion protein S before entry through the plasma membrane, or via a clathrin-dependent endosomal pathway (*18*). MERS-CoV utilizes a similar mechanism but uses the dipeptidyl peptidase 4 receptor (dpp4) instead (*19*). Dpp4 does not share sequence and structural similarity to previously identified human coronavirus receptors such as ACE2 or APN (*16, 17, 20*). Convalescent plasma samples from SARS-CoV-infected patients have moderate cross-reactivity with SARS-CoV-2 spike protein, but no cross-neutralization (*21*). The assay used here has been shown to have no cross-reactivity with the seasonal human coronavirus NL63 (*3*). Interestingly, when we tested human convalescent serum from MERS patients with our ELISA, we found explicit cross-reactivity between MERS serum and SARS-CoV-2 spike with an antibody titer similar to that of SARS-CoV-2 plasma. Our sequence alignment indicated only 18.7 % sequence identity between SARS-CoV-2 RBD and MERS-CoV RBD. On the other hand, we found 32.3 % identity between SARS-CoV-2 and MERS-CoV spike (S) (Supplementary Figure 3), indicating higher sequence conservation in the other domains of the spike protein, thus providing a possible explanation for the observed cross-reactivity. For a given SARS-CoV-2 convalescent serum sample with an anti-spike titer of >1:1350, the probability of viral neutralization at the FDA-recommended level for convalescent plasma used for COVID-19 treatment (viral neutralization titer ≥1:160), has been found to be ≥80 % (*22*). Our data shows strong MERS cross-reactivity with SARS-CoV-2 spike at titers close to 1:1350. Although most effective neutralizing antibodies against coronaviruses target the RBD, neutralizing antibodies against SARS-CoV-2 S1-N-terminal domain (*23*) and SARS-CoV S2 domain (*24*) have also been identified. Our data suggests that MERS convalescent serum may also contain such neutralizing antibodies against SARS-CoV-2.

## Data Availability

Requests for further information or raw data should be directed to the corresponding author.

## Author contributions

M.M., ELISA implementation, validation, analysis. M.M., S.S., N.S., ELISA execution. S.S., test kit development. T.K., N.S., protein expression. T.K., protein purification. T.K., J.H., electron microscopy and image analysis. C.B., workflow automation. K.K., sequence alignment. T.R., S.J., website design, sample codes. M.N. provided finger-prick validation samples, controls, and collected long-term samples. T.M., medical advice and study ethics. M.C., mentoring and advice, website content, study ethics. M.W. project creation and supervision. M.M., M.W. wrote initial draft. All authors edited and contributed to the final manuscript.

## Ethics, consent, and permissions

The experiments were conducted according to a proposal approved by the OIST Human Subjects Research Review Committee (Protocol Title: “Survey of antibody retention rate for OIST staff and students against SARS-CoV-2”; application reference number: HSR-2020-026). All methods, including obtaining informed consent, were conducted in accordance with the Declaration of Helsinki and other relevant guidelines including the Ethical Guidelines for Medical and Health Research Involving Human Subjects set forth by the Japanese government. Consent to publish the accumulated anonymized data has been obtained from all participants of the study upon enrollment. The protocol for the positive control samples has been approved by the Institutional Review Board (IRB) of Okinawa Chubu Hospital with the confirmation number 2020-23. This protocol is furthermore included in the application with reference number HSR-2020-16-2 approved by the OIST Human Subjects Research Review Committee (Protocol Title: “Establishment of new serological diagnostic test by ELISA to grasp the post-infection population with novel coronavirus in Okinawa”).

## Data availability

Original uncropped unscaled images for Figures 1 A-D are available as supplementary files. Requests for further information or raw data should be directed to the corresponding author.

## Acknowledgements

MERS-CoV convalescent sera was kindly provided by Dr Giada Mattiuzzo (NIBSC, United Kingdom), in collaboration with Dr Manki Song (International Vaccine Institute, South Korea) Chungnam National University Hospital, and funded by Coalition for Epidemic Preparedness Innovations (CEPI, Norway). We thank Kieran Deasy and Mouez Lassoued for 3D-printing tube racks. We are grateful to Mahesh Bandi for advice on statistics and to Alejandro Villar Brillones for advice on automation. We thank Chris Wu for developing the website’s theme and stylesheets. We thank Pinaki Chakraborty for providing the time and expertise of his technician C.B. This work was supported by the Platform Project for Supporting Drug Discovery and Life Science Research (BINDS) from AMED, under grant number JP18am0101076 (to M.W.). We acknowledge financial support by OPG for a larger serological study, as well as direct funding from OIST.

