## supplementary files for "COVID-19 serological survey using micro blood sampling"

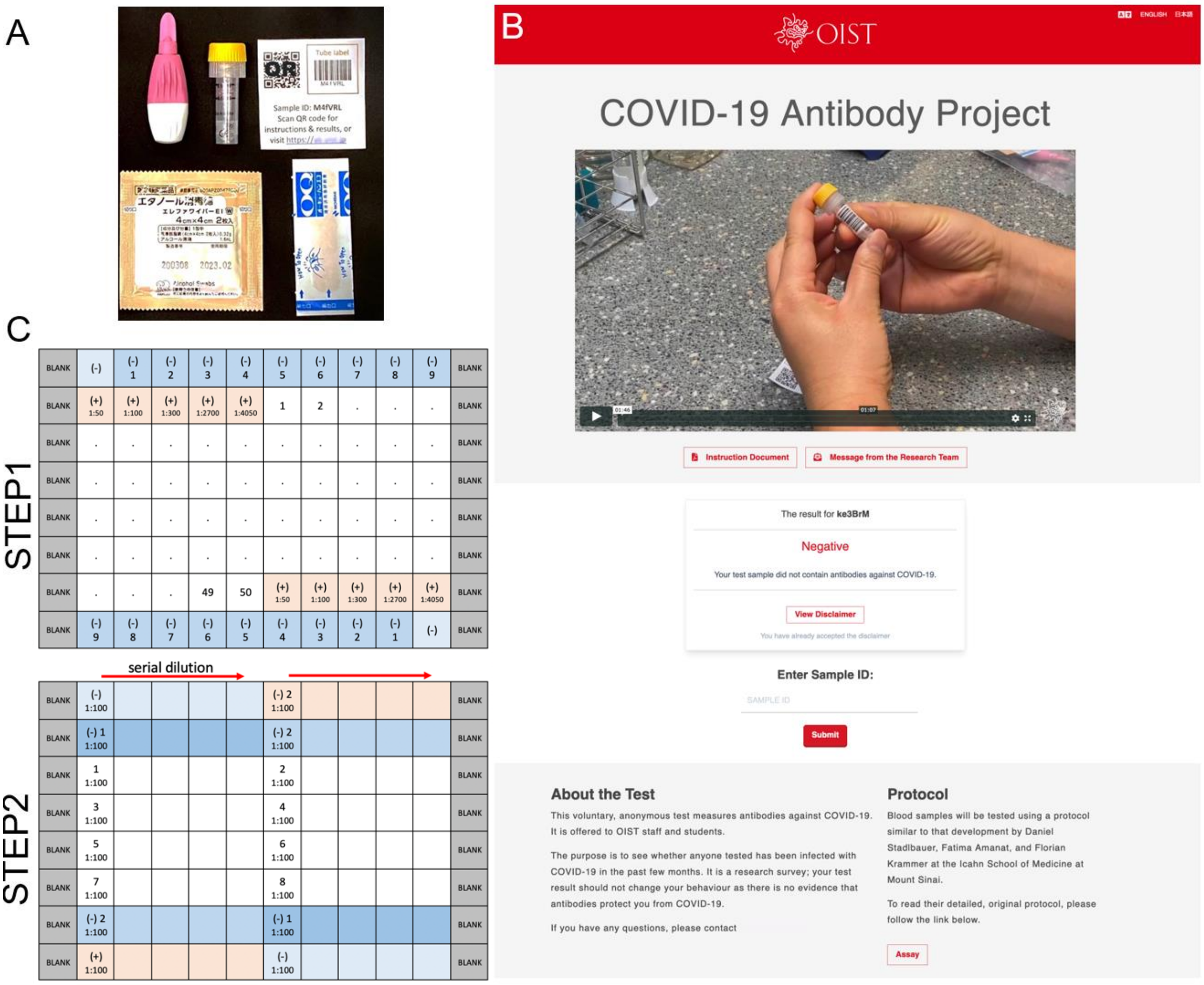


**Supplementary Figure 1: Antibody Survey Sample Collection Kit and Plate Design. A.** Each blood sampling kit included BD Microtainer® contact-activated lancet (Becton Dickison, USA), 0.8 ml volume blood collection tube containing a coagulant and a separator (Greiner bio-one), packaged alcohol wipes, adhesive bandage, and sticker with an identification barcode and QR code. **B.** Front page of OIST Antibody Test website. **C.** Plate designs used in the ELISA. Plates are modified versions of those described previously (*1*).

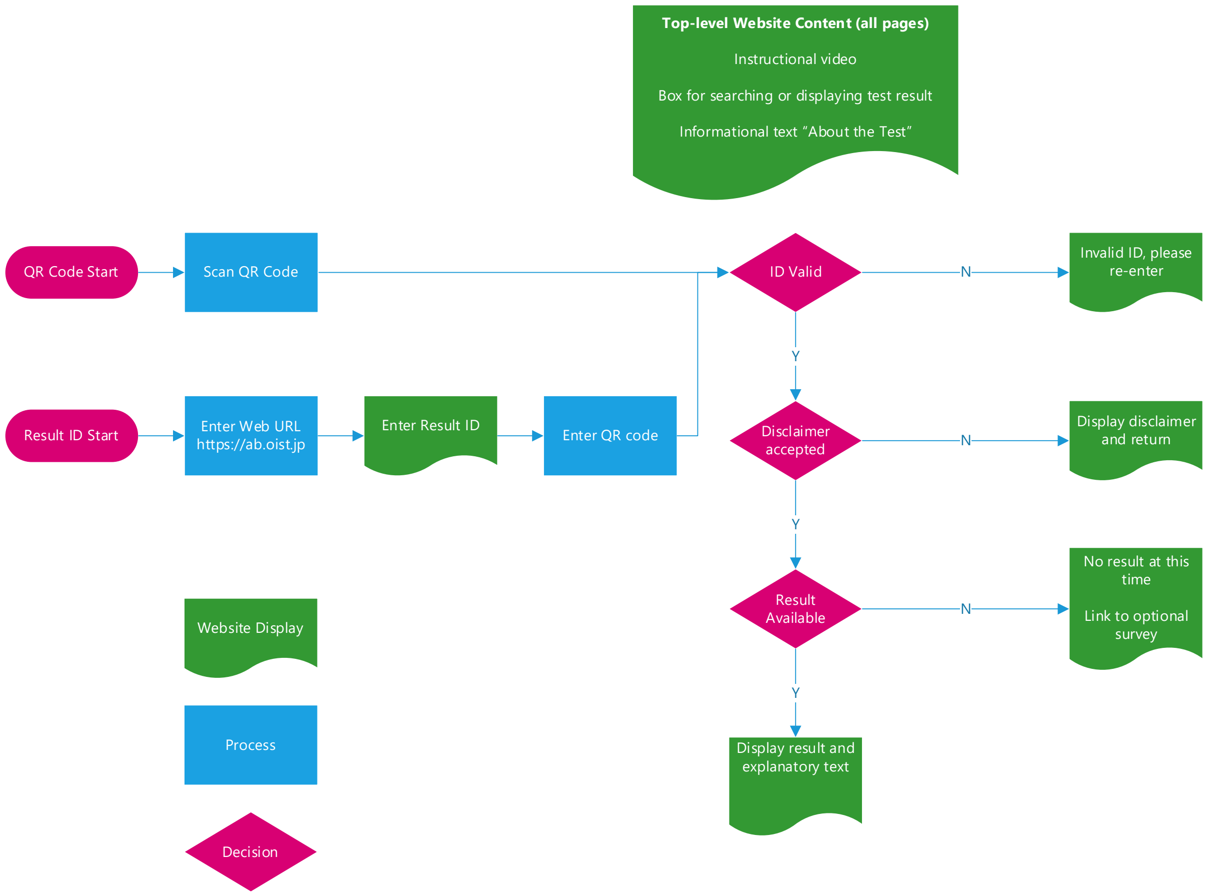


**Supplementary Figure 2:** Basic user flow followed by participants to view test results and answer optional survey. The diagram was created with Microsoft Visio Professional.

```
SARS-CoV-2   ----MFVFLVLL-P----------LVSSQCVNLTTRT-----QLPPAYTNSFTRGVYYPDKVFRSSVLHSTQDLFLPFFSN---VTWFHAIHVSGTNGTKRF----DNPVLPFNDGVYFA  93
MERS         MIHSVFLLMFLLTPTESYVDVGPDSVKSACIEVDIQQTFFDKTWPRPIDVSKADGIIYPQGRTYSNITITYQGLF-PYQGDHGDMYVYSAGHATGTTPQKLFVANYSQDVKQFANGFVVR  119
                 :*:::.** *          *.* *:::  :        *      * : *: **:     *.:  : *.** *: .:    :   : * *.:**.  * *     .: *  * :*. .

SARS-CoV-2   S-------------TEK----SNIIRGWIFGTTLDSKT---------QSLLIVNNATNVVIKVCEFQFCNDPFLGV------------YYHK---------NNKSWMESEFRVYSSANNC  166
MERS         IGAAANSTGTVIISPSTSATIRKIYPAFMLGSSVGNFSDGKMGRFFNHTLVLLPDGCGTLLRA--FYCILEPRSGNHCPAGNSYTSFATYHTPATDCSDGNYNRNASLNSFKEYFNLRNC  237
                       ..      :*  .:::*:::.. :          ::*::: :. ..:::.  *     :*  *              **.           *:.    ..*: * . .**

SARS-CoV-2   TFEYVSQPFLMDLEGKQGNFKNLREFVFKNIDGYFKIYSKHTPINLVR----DLPQGFSALEPLVDLPIGINITRFQTLLALHRSYLTPGDSSSGWTAGAAAYYVGYLQPRTFLLKYNEN  282
MERS         TFMYTYNITE-----------------DEILEWFGITQTAQGVHLFSSRYVDLYGG--NMFQFATLPVYDTIKYYSII---PHSIRSIQSDRKAW----AAFYVYKLQPLTFLLDFSVD  330
             ** *. :                     .:*  :* * ..   ::*.      **  *    :  :. **:  .*. :. :     :*  :   .. ..*    **:**  *** ****.:. :

SARS-CoV-2   GTITDAVDCALDPLSETKCTLKSFTVEKGIYQTSNFRVQPTESIVRFPNITNLCPFGEVFNATRFASVYAWNRKRISNCVADYSVLYNSASFSTFKCYGVSPTKLNDLCFTNVYADSFVI  402
MERS         GYIRRAIDCGFNDLSQLHCSYESFDVESGVYSVSSFEAKPSGSVVEQAE-GVECDFSPLLSGT-PPQVYNFKRLVFTNCNYNLTKLLSLFSVNDFTCSQISPAAIASNCYSSLILDYFSY  448
             * *  *:**.:: **: :*: :** **.*:*..*.*..:*: *:*.   :       * *. ::..*    .** ::*  ::**  : : * .   *.. *.*  :**: : . *::.:  * *

SARS-CoV-2   RGDEVRQIAPGQTGKIADYNYKLPDDFTGCVIAWNSNNLDSKVGGN--YNYLYRLFRKSNLKPFERDISTEIYQAGSTPCNGVEGFNCY----------FPLQSYGFQPTNGVGYQPYRV  510
MERS         PLSMKSDLSVSSAGPISQFNYKQSFSNPTCLILATVPHNLTTITKPLKYSYINKCSR--FLSDDRTEVPQLVNANQYSPCVSIVPSTVWEDGDYYRKQLSPLEGGGWLVASGSTVAMTEQ  566
               .    ::: ..:* *:::***    .    *:*   .   :   :.:       *.*: :   *    *.    . ::     :       :** .:     . :                  **:. *:   :.*       .

SARS-CoV-2   VVLSFEL----LHAPATVCGP-----KKSTNLVKNKCVNFNFNGLTGTGVLTESNKKFLPFQQFGRDIADTTDA-VRDPQTLEILDITPCSFGGVSVITPGTNTSNQVAVLYQDVNCTEV  620
MERS         LQMGFGITVQYGTDTNSVCPKLEFANDTKIASQLGNCVEYSLYGVSGRGVFQNCTAVGVRQQRFVYDAYQNLVGYYSDD--GNYYCLRACVSVPVSVIYD--KETKTHATLFGSVACEHI  682
             : :.* :           :**        ...         .:**::.: *::* **: :..     :   *:*  *  :.   .     *     :     :   *      ****      : ::   *.*: .* * .:

SARS-CoV-2   PVAIHADQL--TPTWRVYSTGSNVFQTRAGCLIGAEHVN-NSYECDIPIGAGICASYQTQTNS-PASVASQ------SIIAYTMSLGAENSVAYSNNSIAIPTNFTISVTTEILPVSMTK  730
MERS         SSTMSQYSRSTRSMLKRRDSTYGPLQTPVGCVLGLVNSSLFVEDCKLPLGQSLCALPDTPSTLTPRSVRSVPGEMRLASIAFNHPIQ-VDQLNSSYFKLSIPTNFSFGVTQEYIQTTIQK  801
               ::     .          :   .:    . :** .**::*    :  .         :*.:*:*  .:**    :* :.    * ** *             : **:.     :     :.:    *    .::*****::.** *  :  .:: *

SARS-CoV-2   TSVDCTMYICGDSTECSNLLLQYGSFCTQLNRALTGIAVEQDKNTQEVFAQVKQIYKTPPIKDFGGFN-FSQILP---DPSKPSKRSFIEDLLFNKVTLADAGFIKQYGDCLG--DIAAR  844
MERS         VTVDCKQYVCNGFQKCEQLLREYGQFCSKINQALHGANLRQDDSVRNLFASVKSSQSSPIIPGFGGDFNLTLLEPVSISTGSRSARSAIEDLLFDKVTIADPGYMQGYDDCMQQGPASAR  921
             .:***. *:*..  :*.:** :**.**::*:** *  :.**...:::**.**.   .:* * .***    :: : *     . ..  * ** ******:***:** *::: *.**:       :**

SARS-CoV-2   DLICAQKFNGLTVLPPLLTDEMIAQYTSALLAGTITSGWTFGAGAALQIPFAMQMAYRFNGIGVTQNVLYENQKLIANQFNSAIGKIQDSLSSTASALGKLQDVVNQNAQALNTLVKQLS  964
MERS         DLICAQYVAGYKVLPPLMDVNMEAAYTSSLLGSIAGVGWTAGLSSFAAIPFAQSIFYRLNGVGITQQVLSENQKLIANKFNQALGAMQTGFTTTNEAFHKVQDAVNNNAQALSKLASELS  1041
             ****** . * .*****:  :* * ***:**..     *** * .:    **** .: **:**:*:**:** ********:**.*:* :* .:::* .*: *:**.**:*****..*..:**

SARS-CoV-2   SNFGAISSVLNDILSRLDKVEAEVQIDRLITGRLQSLQTYVTQQLIRAAEIRASANLAATKMSECVLGQSKRVDFCGKGYHLMSFPQSAPHGVVFLHVTYVPAQEKNFTTAPAICHDGKA  1084
MERS         NTFGAISASIGDIIQRLDVLEQDAQIDRLINGRLTTLNAFVAQQLVRSESAALSAQLAKDKVNECVKAQSKRSGFCGQGTHIVSFVVNAPNGLYFMHVGYYPSNHIEVVSAYGLCDAANP  1161
             ..*****: :.**:.*** :* :.******.*** :*:::*:***:*: .     **:**  *:.*** .**** .***:* *::**  .**:*: *:** * *::. :..:* .:*. .:

SARS-CoV-2   ---HFPREGVFVSN-----GTHWFVTQRNFYEPQIITTDNTFVSGNCDVVIGIVNNTVYDPLQ--PELDSFKEELDKYFKNHTSPDVDLGDISGINASVVNIQKEIDRLNEVAKNLNESL  1194
MERS         TNCIAPVNGYFIKTNNTRIVDEWSYTGSSFYAPEPITSLNTKYVAPQVTYQNI-STNLPPPLLGNSTGIDFQDELDEFFKNVSTSIPNFGSLTQINTTLLDLTYEMLSLQQVVKALNESY  1280
                * :* *:..        .*  *  .** *: **: **    .    .   .* ...:  **        .*::***::*** ::    ::*.:: **::::::  *:   *::*.* ****

SARS-CoV-2   IDLQELGKYEQYIKWP----------------------------------------------------------  1210
MERS         IDLKELGNYTYYNKWPWYIWLGFIAGLVALALCVFFILCCTGCGTNCMGKLKCNRCCDRYEEYDLEPHKVHVH  1353
             ***:***:*  * ***
```

**Supplementary Figure 3:** Amino acid sequence alignment of spike (S) proteins from SARS-CoV-2 and MERS-CoV. RBD regions (SARS-CoV-2, residue 319-541 and MERS, residue 367-606) are highlighted in red. The figure was created with ClustalW2 and reformatted in Adobe Illustrator CC 2020.
